# Understanding the psychological impact of the COVID-19 pandemic and containment measures: an empirical model of stress

**DOI:** 10.1101/2020.05.13.20100313

**Authors:** Bartholomäus Wissmath, Fred W. Mast, Fabian Kraus, David Weibel

## Abstract

Research suggests that epidemics and corresponding containment measures have negative consequences to the individual and cause stress. The psychological mechanisms that determine stress, caused by the COVID-19 pandemic and containment measures, are not yet clear. In a survey during the lockdown in Switzerland (n=1565), we found substantially increased levels of stress in the population. In particular, individuals who did not agree with the containment measures, as well as those who saw nothing positive in the crisis, experienced even higher levels of stress. In contrast, individuals who are part of a risk group or who are working in healthcare or in essential shops experienced similar stress levels as the general public. We conducted a path analysis to gain a deeper understanding of the psychological mechanisms during lockdown. Experiencing fear of the disease is a key driver for being worried. Our model further shows that worries about the individual, social, and economic consequences of the crisis, strongly boost stress. The infection rate in the canton (i.e. state) of residence also contributes to stress. Positive thinking and perceived social, organizational, and governmental support mitigate worries and stress. To prevent stress, authorities should explain containment measures well, highlight positive aspects of the crisis, address worries, and facilitate support.

## Background

COVID-19 evokes stress in patients, healthcare professionals, and relatives (1). The impact may not be limited to those directly affected; health worries and uncertainty are assumed to generate fear, anxiety, and severe stress in the general population (2). Hence, mental health practitioners anticipate a sharp rise in the need for mental health services (3). Some individuals may be more vulnerable to psychosocial consequences; risk factors include mental or physical pre-existing conditions, working in healthcare, or social isolation (4,5).

Switzerland was among the countries first and most affected by COVID-19 in Europe. To curb the spread of the virus, the Swiss government enacted a set of containment measures: encouraging hygiene, giving shelter-in-place orders, as well as closing borders, child-care facilities, schools, restaurants, bars, leisure facilities, and non-essential shops. Events and gatherings of more than five individuals were banned.

These measures directly affect the general population and may come with side effects. Reduced social interactions are risk factors for mental disorders such as major depression (6), and the containment measures may trigger an economic downturn, thus adding further stress (7). Early studies on COVID-19 observed increased stress levels (8). However, little is known about the psychological mechanisms that determine individual stress in this crisis. We assume that fear of COVID-19 and the local infection rate trigger stress (9,10). Individual, economic, and societal worries are also expected to increase stress levels. Agreement with containment measures, perceived support, (9,11), and being optimistic about the crisis (12), are likely to decrease stress. During the lockdown, we surveyed 1565 individuals in Switzerland online.

## Results

First, we compared the observed stress levels in our sample (*M*=15.58; *SD*=6.65) to a representative community sample under normal conditions (*M*=12.57; *SD*=6.24) (13). A one-sample Gauss test showed higher stress levels during the lockdown period, *z*=19.08, p <.001. The effect (d=.48) is medium to strong.

In a second step, we computed multiple comparisons (ANOVA) of different subgroups (see Table 1). Women expressed higher stress levels than men, as has been found in non-epidemic situations (14). Young individuals reported the highest stress levels, whereas the levels were lowest for individuals older than 65 years (small to medium effect). The majority of respondents (66%) agreed with the measures of the Swiss authorities. These individuals scored lower on stress than individuals who felt that the measures were either not sufficient or too extreme. Individuals who strongly disagreed experienced even more stress (medium effect). Individuals who reported that the lockdown situation also has positive aspects (89.4%) had substantially lower stress levels compared to those who did not see any positive aspects at all (strong effect).

**Table 1:**
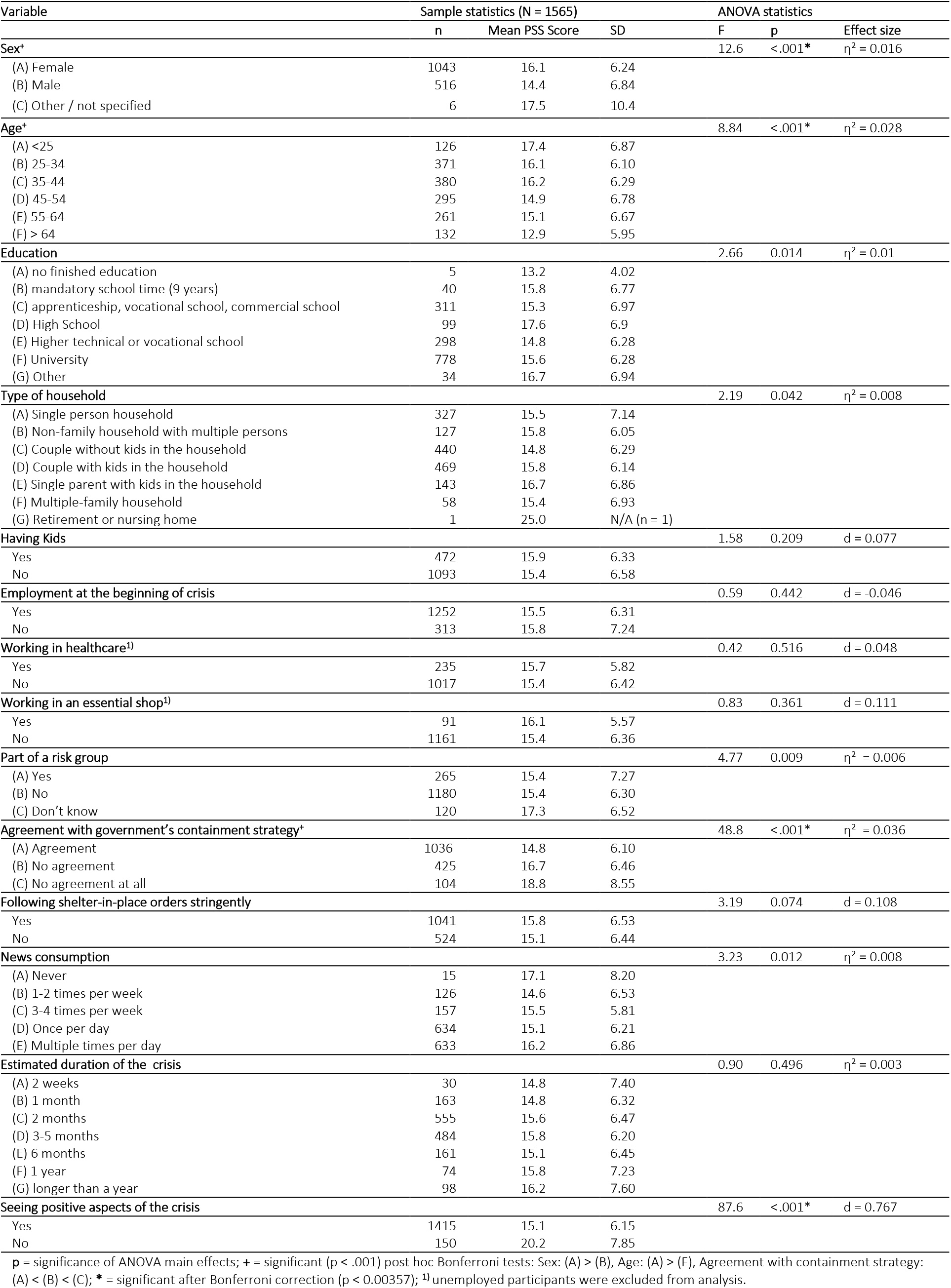
Group comparisons of experienced stress

| Variable | Sample statistics (N = 1565) |  |  | ANOVA statistics |  |  |
| --- | --- | --- | --- | --- | --- | --- |
|  | n | Mean PSS Score | SD | F | p | Effect size |
| **Sex\*** | | | | 12.6 | <.001* | $\eta^2 = 0.016$ |
| (A) Female | 1043 | 16.1 | 6.24 |  |  |  |
| (B) Male | 516 | 14.4 | 6.84 |  |  |  |
| (C) Other / not specified | 6 | 17.5 | 10.4 |  |  |  |
| **Age\*** | | | | 8.84 | <.001* | $\eta^2 = 0.028$ |
| (A) <25 | 126 | 17.4 | 6.87 |  |  |  |
| (B) 25-34 | 371 | 16.1 | 6.10 |  |  |  |
| (C) 35-44 | 380 | 16.2 | 6.29 |  |  |  |
| (D) 45-54 | 295 | 14.9 | 6.78 |  |  |  |
| (E) 55-64 | 261 | 15.1 | 6.67 |  |  |  |
| (F) > 64 | 132 | 12.9 | 5.95 |  |  |  |
| **Education** | | | | 2.66 | 0.014 | $\eta^2 = 0.01$ |
| (A) no finished education | 5 | 13.2 | 4.02 |  |  |  |
| (B) mandatory school time (9 years) | 40 | 15.8 | 6.77 |  |  |  |
| (C) apprenticeship, vocational school, commercial school | 311 | 15.3 | 6.97 |  |  |  |
| (D) High School | 99 | 17.6 | 6.9 |  |  |  |
| (E) Higher technical or vocational school | 298 | 14.8 | 6.28 |  |  |  |
| (F) University | 778 | 15.6 | 6.28 |  |  |  |
| (G) Other | 34 | 16.7 | 6.94 |  |  |  |
| **Type of household** | | | | 2.19 | 0.042 | $\eta^2 = 0.008$ |
| (A) Single person household | 327 | 15.5 | 7.14 |  |  |  |
| (B) Non-family household with multiple persons | 127 | 15.8 | 6.05 |  |  |  |
| (C) Couple without kids in the household | 440 | 14.8 | 6.29 |  |  |  |
| (D) Couple with kids in the household | 469 | 15.8 | 6.14 |  |  |  |
| (E) Single parent with kids in the household | 143 | 16.7 | 6.86 |  |  |  |
| (F) Multiple-family household | 58 | 15.4 | 6.93 |  |  |  |
| (G) Retirement or nursing home | 1 | 25.0 | N/A (n = 1) |  |  |  |
| **Having Kids** |  |  |  | 1.58 | 0.209 | d = 0.077 |
| Yes | 472 | 15.9 | 6.33 |  |  |  |
| No | 1093 | 15.4 | 6.58 |  |  |  |
| **Employment at the beginning of crisis** |  |  |  | 0.59 | 0.442 | d = -0.046 |
| Yes | 1252 | 15.5 | 6.31 |  |  |  |
| No | 313 | 15.8 | 7.24 |  |  |  |
| **Working in healthcare<sup>1)</sup>** |  |  |  | 0.42 | 0.516 | d = 0.048 |
| Yes | 235 | 15.7 | 5.82 |  |  |  |
| No | 1017 | 15.4 | 6.42 |  |  |  |
| **Working in an essential shop<sup>1)</sup>** |  |  |  | 0.83 | 0.361 | d = 0.111 |
| Yes | 91 | 16.1 | 5.57 |  |  |  |
| No | 1161 | 15.4 | 6.36 |  |  |  |
| **Part of a risk group** | | | | 4.77 | 0.009 | $\eta^2 = 0.006$ |
| (A) Yes | 265 | 15.4 | 7.27 |  |  |  |
| (B) No | 1180 | 15.4 | 6.30 |  |  |  |
| (C) Don't know | 120 | 17.3 | 6.52 |  |  |  |
| **Agreement with government's containment strategy\*** | | | | 48.8 | <.001* | $\eta^2 = 0.036$ |
| (A) Agreement | 1036 | 14.8 | 6.10 |  |  |  |
| (B) No agreement | 425 | 16.7 | 6.46 |  |  |  |
| (C) No agreement at all | 104 | 18.8 | 8.55 |  |  |  |
| **Following shelter-in-place orders stringently** |  |  |  | 3.19 | 0.074 | d = 0.108 |
| Yes | 1041 | 15.8 | 6.53 |  |  |  |
| No | 524 | 15.1 | 6.44 |  |  |  |
| **News consumption** | | | | 3.23 | 0.012 | $\eta^2 = 0.008$ |
| (A) Never | 15 | 17.1 | 8.20 |  |  |  |
| (B) 1-2 times per week | 126 | 14.6 | 6.53 |  |  |  |
| (C) 3-4 times per week | 157 | 15.5 | 5.81 |  |  |  |
| (D) Once per day | 634 | 15.1 | 6.21 |  |  |  |
| (E) Multiple times per day | 633 | 16.2 | 6.86 |  |  |  |
| **Estimated duration of the crisis** | | | | 0.90 | 0.496 | $\eta^2 = 0.003$ |
| (A) 2 weeks | 30 | 14.8 | 7.40 |  |  |  |
| (B) 1 month | 163 | 14.8 | 6.32 |  |  |  |
| (C) 2 months | 555 | 15.6 | 6.47 |  |  |  |
| (D) 3-5 months | 484 | 15.8 | 6.20 |  |  |  |
| (E) 6 months | 161 | 15.1 | 6.45 |  |  |  |
| (F) 1 year | 74 | 15.8 | 7.23 |  |  |  |
| (G) longer than a year | 98 | 16.2 | 7.60 |  |  |  |
| **Seeing positive aspects of the crisis** |  |  |  | 87.6 | <.001* | d = 0.767 |
| Yes | 1415 | 15.1 | 6.15 |  |  |  |
| No | 150 | 20.2 | 7.85 |  |  |  |

p = significance of ANOVA main effects; + = significant ( $p < .001$ ) post hoc Bonferroni tests: Sex: (A) > (B), Age: (A) > (F), Agreement with containment strategy: (A) < (B) < (C); \* = significant after Bonferroni correction ( $p < 0.00357$ ); <sup>1)</sup> unemployed participants were excluded from analysis.

No other assessed variable influenced stress (cf. Table 1). Interestingly, individuals identifying as members of a risk group, as well as individuals working in healthcare or in essential shops with customer contact, did not experience more stress than the rest of the population. Also, the expected duration of the pandemic and amount of news consumption had no influence on stress.

In a third step, we conducted a path analysis with stress as the target variable. Figure 1 depicts the resulting model, showing a good fit.

**Figure 1.**
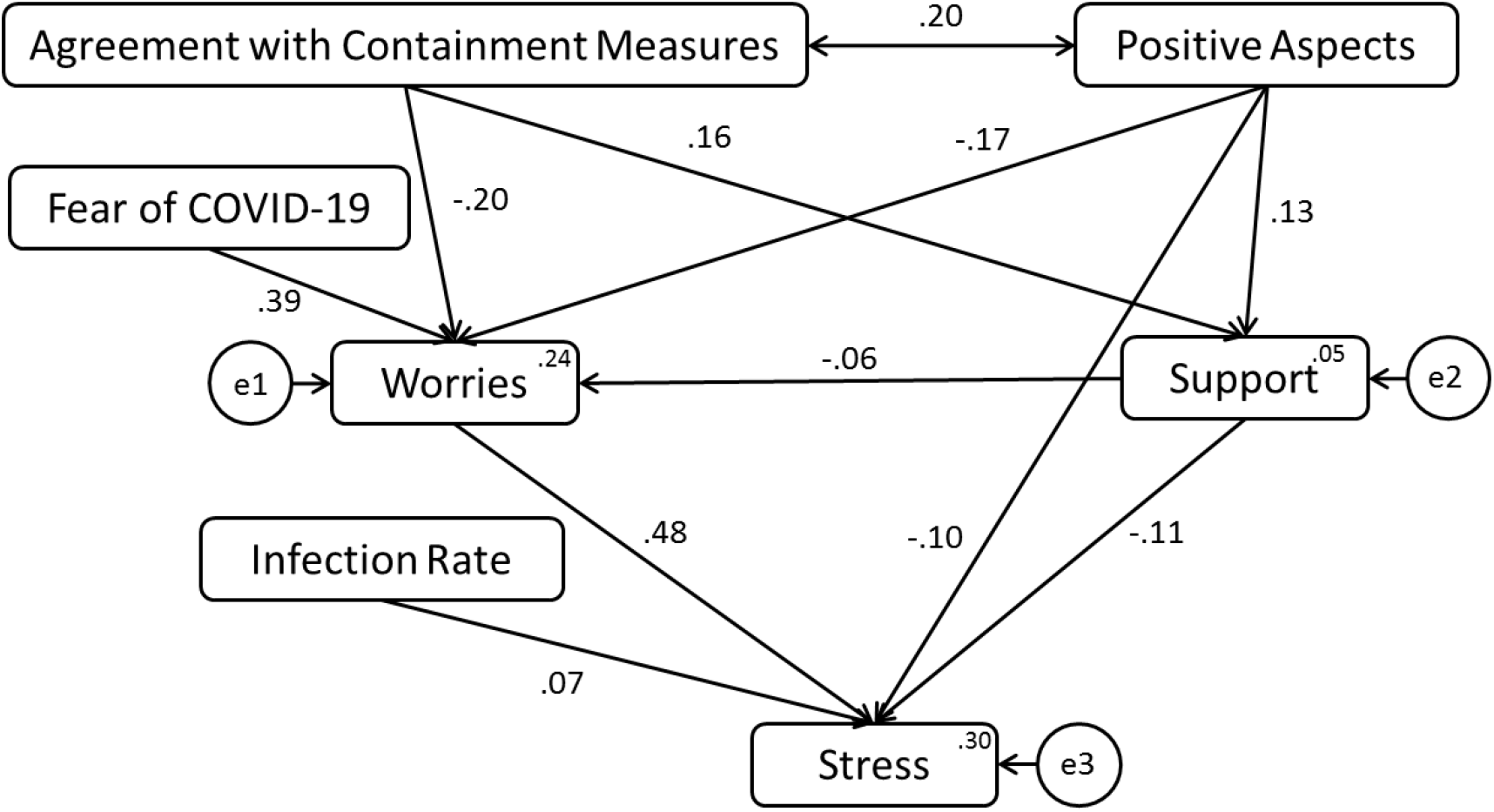
A path model showing the influence of various determinants on stress. Model fit statistics: χ^2^ (10)=42.85, p <.01, GFI=.99, CFI=.97, NFI=.96, TLI=.94, RMSEA=.05. All displayed paths are highly significant (p <.01.). Error variances appear in small circles.

In total, the measured variables explain 30% of variance in the endogenous variable, stress. Worries contribute the most to stress, and worries mediate between fear of COVID-19 and stress. Seeing positive aspects of the crisis, as well as agreement with the government’s containment strategy, are correlated, and both mitigate worries and boost perceived support. In addition, positive thinking directly minimizes stress. Perceived support from family, friends, organizations, and authorities mitigates worries and reduces stress.

## Conclusion

Our results show that the COVID-19 pandemic and the mitigation strategies increase the population’s stress level. In our model, fear of the virus is the most important booster of worries and the amount of worries is the most important driver of stress. Agreeing with the authorities’ containment strategy and seeing positive aspects of the crisis are important factors mitigating stress. Correspondingly, feeling that the containment measures are not sufficient or too extreme is associated with more stress. Support provided by family, friends, organizations, and authorities are also important protective factors for stress.

In our study, risk factors for high stress levels are a high amount of worries and perceived lack of support and, to a lesser extent, high infection rates. Eleven percent of the respondents do not see anything positive about the crisis. This group experiences substantially more stress than individuals who see at least one positive aspect. Although seeing something positive about the crisis may be associated with the individual situation, personality traits, and respondent’s coping behavior (12), our findings indicate that recognizing positive aspects alleviates stress. Our model builds upon existing theories of stress and explains 30% of the variance in stress.

Surprisingly, members of the risk groups, old respondents and individuals working in healthcare or essential shops with customer contact, did not score higher on stress. One reason could be that extensive protection measures had been mandated. In addition, healthcare providers in Switzerland have so far not been overwhelmed by cases of COVID-19.

Our model could be useful in understanding and addressing the psychological impact of possible new waves of COVID-19 cases and other epidemics. Mitigation measures boost worries and stress, particularly for those individuals who feel that these measures are not sufficient or go too far. Highlighting positive aspects about the crisis and convincing people of the effectiveness and the necessity of containment measures may not only boost compliance, but also decrease stress, since individuals feel protected by the authorities and experience less worries. This, in turn, will hopefully limit the impact on mental health as a consequence of the crisis. Providing support is another important way to mitigate worries, enable coping, and reduce stress.

## Materials and Methods

### Participants

From March 27 to April 26, 2020, a total of 1565 individuals completed the online survey. They were recruited via academic institutions and social media. All participants were Swiss residents. Information about the sample is provided in Table 1. Individuals from Geman-speaking (n=1206), French-speaking (n=247) and Italian-speaking (n=112) regions, were surveyed. The study was reviewed and approved by the Ethics Committee of the Faculty of Human Sciences, University of Bern.

### Measurements

#### Stress

We used the Perceived Stress Scale (PPS-10) (10-items) (13, 14). The widely-used PSS is based on the stress-related components of unpredictable, uncontrollable, and overloading life events (e.g. “In the last month, how often have you been angry because of things that were outside your control?”), (0=never; 4=very often). A PSS score was calculated by summing up all items, with a high score indicating high stress levels. Reliability was high (Chronbach’s alpha=.86).

#### Amount of worries

Participants evaluated to what extent they were worried about the following aspects (1=not at all; 5=a lot): physical health, mental health, health of family and friends, security, social life, personal needs, financial situation, job security, economic situation, healthcare, and basic supply. A principal component analysis on these items suggest a one-factorial solution. For our analyses, we thus computed the mean value across all items.

#### Fear of COVID-19

Fear was measured with one single item: “How afraid are you of the coronavirus (COVID-19)?” (1=not at all; 5=very much).

#### Support

Participants were asked to what extent they felt supported by the following individuals, organizations, or authorities: family, friends, neighbors, employers, authorities, schools, day-care centers, church, primary care physician, hospitals (0=little or no support; 3=strong support). To compute the score, we summed up the ratings.

#### Infection rate

The cases per 100,000 inhabitants in the participants’ resident canton (state) were used (retrieved on April 30 2020: (https://www.corona-data.ch/).

#### Agreement with containment measures

We asked the participants whether they thought that the measures were (a) not at all strict enough (b) not strict enough (c) just right (d) too strict (e) much too strict. For the analyses, the categories (a) and (e) were combined as “no agreement at all,” and the categories (b) and (d) were combined as “no agreement.” These combined categories were contrasted with (c) “agreement.”

#### Seeing positive aspects

We asked the respondents whether there were any positive aspects for them in the current situation (1=No; 2=Yes).

#### Participants’ individual situation and demographics

We assessed the following: sex, age, education, type of household, having kids, employment, working in healthcare or essential shops with customer interaction, part of a risk group, media consumption, and estimated length of the crisis.

## Data Availability

Replication data are available on the Open Science Framework.

https://osf.io/grvwa/

## Data Availability

Replication data are available on the Open Science Framework (15), https://osf.io/grvwa/

## Notes

### Competing Interest Statement

The authors have declared no competing interest.

### Funding Statement

No external funding was received.

